# Long-term and low-level envelope C2V3 stimulation from highly diverse virus isolates leads to frequent development of broad and elite antibody neutralization in HIV-1 infected individuals

**DOI:** 10.1101/2022.01.27.22269759

**Authors:** Francisco Martin, José Maria Marcelino, Claudia Palladino, Inês Bártolo, Susana Tracana, Inês Moranguinho, Paloma Gonçalves, Rita Mateus, Rita Calado, Pedro Borrego, Thomas Leitner, Sofia Clemente, Nuno Taveira

**Author notes:** Address correspondence to: Nuno Taveira Research Institute for Medicines (iMed.ULisboa), Faculty of Pharmacy, Universidade de Lisboa Av. Prof. Gama Pinto, 1649-003 Lisbon, Portugal.

## Abstract

Elicitation of potent neutralizing antibodies against genetically diverse HIV-1 isolates is important for an effective HIV-1 vaccine. Some HIV-1 infected patients produce such broadly neutralizing antibodies (bNAbs). Identification of host and viral correlates of bNAb production may help develop the next generation of HIV-1 vaccines. We carried out the first detailed characterization of the neutralizing antibody response and identify viral and host factors associated with the development of bNAbs in HIV-1 infected patients from Angola, one of the oldest, more dynamic, and diverse HIV-1 epidemics in the world. Plasma samples from 322 HIV-1 infected patients were collected in 2001, 2009 and 2014. Phylogenetic analysis of C2V3C3 envelope sequences identified a diverse array of subtypes including A1, A2, B, C, D, F1, G, H, J, untypable strains, and recombinant forms which prevailed over pure subtypes. Notably, 56% of the patients developed cross, broad, or elite neutralizing responses against a reference panel of tier 2 Env-pseudoviruses far exceeding results obtained elsewhere in the world. The frequency of elite neutralizers was higher in 2014, when patients were on ART and had low viremia, than in 2009 when patients were drug naive. In drug naïve patients, broad neutralization was associated with subtype C infection, lower CD4+ T cell counts, higher age, or higher titer of C2V3C3-specific antibodies relative to patients that did not develop bNAbs. Neutralizing antibodies targeted the V3-glycan supersite in most patients but antibodies specific for the V2 apex, the CD4 binding site, the gp41 membrane-proximal external region (MPER) and unknown epitopes were also found in some patients. V3 and C3 regions were significantly less variable and less subject to positive selection in elite neutralizers compared to weak or no neutralizers suggesting an active role of bNabs directed against these regions in controlling HIV-1 replication and diversification. Hence, development of broad and elite antibody neutralization against HIV-1 requires long-term and low-level envelope V3C3 stimulation from highly diverse subtype C isolates. These results have direct implications for the design of a new generation of HIV-1 vaccines.

## Introduction

The HIV-1 Env glycoprotein is highly immunogenic but, in general, the antibodies elicited by it during infection lack neutralizing breadth or potency against primary HIV-1 strains thus failing to inhibit viral replication in infected individuals ^1, 2^. In natural infection only 5% to 30% of adults develop broadly neutralizing antibodies (bNAbs) after several years of infection ^2, 3, 4, 5, 6, 7^ and these bNAbs have little impact in the control of the infection due to the continuous capacity of HIV-1 to diversify and escape these antibodies ^5, 8, 9^. However, some recombinant human bNAbs supress viral replication in HIV-1 infected individuals ^10, 11, 12, 13, 14, 15^, prevent human infection by some HIV-1 strains ^16^, and passive immunization in animal models can protect from infection and/or disease progression (reviewed in ^17^). Therefore, bNAbs are promising tools to restrict HIV-1 transmission and control disease progression if they could be induced by vaccination. Unfortunately, so far, antibodies elicited by candidate immunogens and vaccines have shown a limited ability to neutralize heterologous primary HIV-1 strains ^8, 18, 19, 20, 21, 22, 23, 24^.

bNAbs target five highly conserved epitopes in the HIV-1 envelope: the CD4 binding site (CD4bs); the V2 apex; V3 glycan supersite; gp41 MPER, and gp41/gp120 interface which includes the fusion peptide ^17, 25, 26^. However, the mechanisms underlying the elicitation of such antibodies by B cell populations are still largely unknown ^27, 28, 29, 30^. Guiding the immune system to elicit such bNAbs remains a major challenge due to the extremely complex antibody maturation pathways and high levels of somatic hypermutation required by HIV-1 specific antibodies to acquire neutralization breadth ^30, 31^. An exception is the V3-glycan supersite bNAb lineage that does not require extensive antibody-affinity maturation ^32, 33^ allowing their development in early stages of infection ^34^, and explaining their high prevalence in recently infected individuals ^3^. Furthermore, V3-specific IgG binding and neutralizing responses in pregnant woman living with HIV-1 predict low risk of mother-to-child-transmission of the virus ^35^. In HIV-2 infected individuals, the V3 loop is a dominant target of bNAbs such that V3 undergoes extensive sequence, conformational and functional alterations to escape antibody neutralization ^36, 37, 38^. Such findings, together with the proved therapeutic value of V3-glycan supersite bNAbs ^12^, highlight this epitope as a key target for HIV vaccine design.

Understanding the mechanisms underlying the production of bNAbs against HIV-1 in some individuals during natural infection is of crucial importance for the development of improved immunogens and immunization strategies. Gray et al. ^39^ showed that patients infected with HIV-1 clade C rarely produce antibodies binding to the 2F5 neutralizing epitope in gp41 suggesting a correlation between HIV-1 subtype and neutralizing response. However, other studies found limited to no impact of HIV-1 subtype in plasma neutralization, suggesting that HIV-1 group M subtypes and neutralization response evolved independently ^5, 40, 41^. More recently, in a large longitudinal Sub-Saharan HIV primary infection cohort, cross-clade plasma neutralization was strongly correlated with subtype C infection ^3^. Additionally, Rusert et al. ^2^ found a strong association between plasma neutralization specificity and HIV-1 subtype, with subtype B viruses being more vulnerable to CD4-binding-site specific antibodies and non-B viruses being more vulnerable to V2-glycan specific neutralizing antibodies. In this study, V3-glycan and MPER-specific neutralizing responses were independent of viral subtype. The differences observed between studies might be related with the different assay conditions used to assess neutralizing activity, in particular with the selected indicator virus panel that should represent the global HIV-1 diversity and be standardized to allow inter-study comparison ^42^.

Considering that vaccine effectiveness will depend on the extent to which induced antibodies will neutralize the global diversity of circulating HIV-1 variants, it is important to characterize HIV-1 antibody responses in different epidemics and geographies. For example, we recently showed that the frequency and level of antibody response to selected epitopes in the envelope gp41 differ between HIV-1 infected patients from Germany, France, and Portugal which have different subtype distribution ^43^. In Switzerland, data analysis of the Swiss HIV Cohort reported that ethnicity was associated with bnAb induction being black participants more prone to develop bNAb responses ^2^.

The neutralizing antibody response of HIV-1 infected patients from Angola has never been evaluated. Angolan HIV-1 epidemic is peculiar, as it is driven by all subtypes, multiple circulatory recombinant forms (CRFs) and unique recombinant forms (URFs) ^44, 45, 46, 47^. In addition, being a very old epidemic, highly divergent and ancestral forms of the different subtypes are often present ^45, 47, 48, 49^. It has been suggested that the genetic complexity of the virus quasispecies present in HIV-1 individuals is directly related to the development of neutralization breadth regardless of infection duration (reviewed in ^50^). This should be particularly evident in old epidemics such as the one of Angola. Hence, characterizing the antibody responses and HIV-1 evolution in this population may provide new insights into the development and evolution of the neutralizing antibody response against HIV-1 and into vaccine design. Here, we carried out the first detailed characterization of the neutralizing antibody response against HIV-1 in Angola and identified viral and host factors associated with the neutralizing response.

## Materials and Methods

### Study population and ethics statement

This cross-sectional retrospective study included 322 HIV-1 infected adults. Plasma samples were collected in 2001, 2009 and 2014 at the Hospital da Divina Providência (HDP), a referral hospital in Luanda, the capital city of Angola. Eligible participants had ≥19 years of age, were not pregnant and had a serological diagnosis of HIV-1 [Determine HIV-1/2 (Abbott) and Uni-Gold Recombigen (Trinity Biotech) rapid tests]. Plasma viral load and number of CD4+ T cells were determined in a subset of patients using the Abbott Real Time HIV-1 assay (Abbott Laboratories) and the ABACUS 5 Junior Hematology analyser, respectively. The study was conducted according to the Declaration of Helsinki and was reviewed and approved by the National Ethics Committee of Angola. The study was verbally explained to all the patients before obtaining their written consent.

### Cell lines

TZM-bl and HEK-293T cell lines were obtained from the NIH AIDS Reagent Program (https://www.niaid.nih.gov/research/nih-aids-reagent-program). TZM-bl cells were engineered from HeLa cells that constitutively express CXCR4 to express large amounts of CD4, CCR5 and a firefly luciferase reporter gene under the control of the HIV-1 LTR ^51^. Cells were cultured at 37°C, 5% CO_2_ using Dulbecco minimal essential medium (DMEM) supplemented with 10 % heat-inactivated fetal bovine serum and with 100 units/ml of penicillin and 100 µg/ml of streptomycin.

### Viral RNA extraction, PCR amplification, sequencing, and phylogenetic analysis

Before viral RNA extraction, 1 ml of plasma was centrifuged at 35,000rpm (61,793g) for 1h at 4°C to concentrate viral particles. Supernatant was stored at -80°C for other applications and pelleted material was resuspended with 560 μl of Buffer AVL+RNA carrier from the QIAmp® Viral RNA Mini Kit (Qiagen) and the manufacturer’s protocol was followed. Reverse transcription was performed with NZY First-Strand cDNA Synthesis Kit (NZYtech, Portugal) and a 534bp fragment comprising the C2V3C3 *env* region was amplified by PCR using an in-house method described elsewhere ^47, 52^. Sequencing of the C2V3C3 amplicons was performed with BigDye Terminator Cycle Sequencing Kit (Applied Biosystems). Sequences were aligned with reference strains collected from the Los Alamos HIV Sequence Database (http://www.hiv.lanl.gov/) using Muscle in MEGA version 6 software ^53^. Maximum-likelihood (ML) phylogenetic analyses were performed using the best-fit model of nucleotide substitution as estimated by Modeltest v3.7 under the Akaike information criterion ^54^. ML trees were inferred with PhyML 3.0 ^55^. Tree searching was done with nearest neighbour interchange and subtree pruning and regrafting. The reliability of the obtained topology was estimated with bootstrap with 500 replicates ^55^.

Determination of coreceptor usage was made based on the V3 loop sequence using geno2pheno [coreceptor] webtool (https://coreceptor.geno2pheno.org/) ^56^. False positive rates (FPR) were 10% as recommended ^57^.

Selective pressure was examined with the DATAMONKEY web-server (https://www.datamonkey.org/) ^58^, after removing all positions containing gaps and missing data from the dataset. All estimations were performed using the MG94 codon substitution model crossed with the nucleotide substitution model GTR previously selected with Modeltest. Four different approaches were used to identify codons under selection: single-likelihood ancestor counting (SLAC), fixed-effects likelihood (FEL), internal fixed effects likelihood (IFEL) and relaxed-effects likelihood (REL) methods ^59^. While SLAC, FEL and REL detect sites under selection at the external branches of the phylogenetic three, IFEL identifies such sites only along the internal branches. To classify a site as positively or negatively selected the cut-off P-value was 10% for SLAC, FEL and IFEL. For REL, codons under selection were detected with a cut-off value for the Bayes factor of 50.

### Entropy and N-linked glycosylation analysis

Potential N-linked glycosylation sites were identified using the N-Glycosite software ^60^, and the entropy at each amino acid position was measured with Shannon’s entropy-one and Shannon’s entropy-two online tools, all available at the Los Alamos National Laboratory HIV sequence database (http://www.hiv.lanl.gov/).

### Production of C2V3C3 polypeptides and analysis of antibody reactivity

Six 178 amino acids long polypeptides comprising part of C2, V3 and part of C3 envelope regions (position 212–390 in gp120 in HIV-1 HXB2) of HIV-1 isolates circulating in Angola (subtypes C, G, H, J, and CRF02_AG) and Portugal (subtype B) were expressed in *Escherichia coli* and purified as described previously ^61^. Briefly, a DNA fragment of 534 nucleotides comprising the C2, V3 and C3 coding regions (position 6858-7392 in HIV-1 HXB2) was amplified from plasmids containing the full-length envelope gene using the primers described elsewhere and cloned into the bacterial expression vector pTrcHis (Invitrogen)^61^. Expression of C2V3C3 polypeptides in *Escherichia coli* strain TOP10 was induced with isopropyl-β-D-thiogalactopyranoside (IPTG) according to the manufactureŕs instructions, and protein purification was performed using Dynabeads® His-tag Isolation & Pulldown (Life Technologies). Bradford assay (Bio-Rad) was performed to determine protein concentration. Purified recombinant polypeptides were analysed by SDS-12% PAGE.

Antibody reactivity against these polypeptides was determined using an ELISA assay as described ^61^. In brief, 96-wells ELISA plates were coated overnight at 4^0^C with 100 μl of 0.05M bicarbonate-coating buffer (pH=9.4) containing 100 ng of the different C2V3C3 polypeptides. Plates were blocked with 2% gelatine in Tris-buffered saline (TBS) 1X for 1h at room temperature. Serial dilutions of plasma (1:100 to 1:3200) in primary antibody buffer (TBS 1X + 0.05 % Tween 20 + 5 % blocking solution) were added to the wells and the plates were incubated for 2h at room temperature. Goat anti-human IgG conjugated to alkaline phosphatase, diluted 1:2000 in the primary antibody buffer and 5% goat serum was added to the wells. Plates were washed between steps with TBSt (TBS 1X + 0.05 % Tween 20). Plates were developed by adding Sigma-fast p-nitrophenol phosphate diluted in deionized water. The plates were incubated for 20 minutes at room temperature protected from light. Optical density (OD) was read at 405nm on a microplate reader. The clinical cut-off value of the assay was calculated as the mean OD value of HIV-seronegative samples plus two times the standard deviation (SD). Binding antibody titers were calculated as the highest plasma dilutions giving a positive reaction (OD / cut-off> 1).

### Production of Env-pseudotyped viruses

A reference panel of 12 tier 2 HIV-1 Env-pseudotyped viruses of subtypes C (n=3), A (n=1), CRF07_BC (n=2), CRF01_AE (n=2), B (n=2), G (n=1) and AC recombinant (n=1) were produced using the Global panel of HIV-1 Env clones ^42^, obtained through the NIH AIDS reagent program. Env-pseudotyped viruses were produced by transfection of Env-expressing plasmids in 293T cells using pSG3.1Δenv as backbone, in a 1:3 ratio using JetPRIME® DNA transfection reagent. Viral stocks were filtered through 0.45 µm pore size filters after 48 hours and stored at -80°C until use.

### Plasma neutralization assay

Neutralization of the Env-pseudotyped viruses was assessed in TZM-bl cells using Tat-induced luciferase (Luc) reporter gene expression to quantify the reduction in virus infection as described previously ^62^. Briefly, TZM-bl cells (10,000 cells/well) were seeded the day before the neutralization assay to allow adherence of the cells to the bottom of the wells. Heat inactivated plasma samples (56^0^C for 30 min) were incubated at 1:40 dilution in triplicate with the respective Env-pseudotyped virus for 1 hr at 37^0^C before transfer to TZM-bl cells. After 48 hours, percent neutralization was determined by calculating the difference in average relative light units (RLU) between test wells containing plasma samples and the wells containing the Env-pseudotyped virus from the indicator panel after the normalization of the results using the average RLU of cell control wells. Results were considered valid if the average RLU of virus wells was >10 times the average RLU of cell control wells. A virus pseudotyped with the envelope glycoprotein of vesicular stomatitis virus (VSV-G) was used as neutralization specificity control.

Neutralizing antibody titers were determined for a subset of plasma samples with known antibody profiles (n=64). In this case, 100 µL of 2-fold serial dilutions beginning at 1:40 were mixed with 100 µL of each virus (200 TCID50/well) and incubated for 1 h before adding to the cells. After 48 h, culture medium was removed from each well, and plates were analyzed for luciferase activity as described above. Wells with medium were used as background control, and virus-cell wells were included as infection control. Neutralizing titer (ID50) was defined as the highest dilution for which 50% neutralization was achieved.

### Neutralization score and plasma categorization

To categorize the neutralizing activity of the Angolan samples in terms of potency and breadth we used a previously described scoring system ^2, 43, 63^. A score of 0 was attributed when neutralizing activity against a given virus of the panel was less than 20%, a score of 1 when neutralization ranged between 20 and <50%, a score of 2 for 50 to <80% neutralization and a score of 3 for ≥80% neutralization. The overall neutralization score (NS) for a given plasma was obtained by adding the scores against the 12 Env-pseudoviruses of the panel and reflects neutralization potency and breadth. As a validated and worldwide accepted classification system to define neutralizing activity is lacking, for the purpose of the present study we classified plasmas with scores 25-36 as elite neutralizers, 18-24 as broad neutralizers, 6-17 as cross neutralizers and <6 as weak or no neutralizers. According to this classification a plasma sample from an elite neutralizer must neutralize ≥9 viruses of the panel with a neutralization potency ≥80%.

### Prediction of bNAb epitope specificities by clustering analysis

The neutralizing antibody specificities were determined for a subset of patients exhibiting broad and elite neutralization capacity using clustering analysis with human bNAbs targeting the main neutralizing epitopes on the viral envelope and capable to neutralize at least half of the 12 Env-pseudotyped viruses of the panel, as described previously ^42^. Neutralization heatmaps and clusters were computed via the online tool ClustVis using a predefined correlation clustering distance method (Pearson correlation subtracted from 1) based on the average distance of all possible pairs. ClustVis is a web tool for visualizing clustering of multivariate data (available at https://biit.cs.ut.ee/clustvis/) ^64^.

### Statistical analysis

Statistical analysis was performed with GraphPad Prism versions 5.01 or 9.0 (GraphPad Software Incorporated, San Diego, California, USA). The Mann-Whitney, Kruskal-Wallis or Fisher’s exact tests were used to compare differences between groups. The Spearman rank test was used to quantify the magnitude and direction of the correlation between antibody neutralization activity and plasma binding titers against C2V3C3-polypeptides, CD4+ T cell counts, viral subtype and age of patients. Hypothesis tests were two-tailed and P values <0.05 were considered significant. To test the potential correlation between neutralization score and genetic distance of the clinical samples to the neutralization panel viruses, we used amino acid sequences and Hamming distances that included gaps as characters because: 1) neutralization occurs on the amino acid level, 2) does not depend on the evolutionary path to a state-combination, and 3) indels may have significant effects on antibody binding. Genetic distances were calculated using DECIPHER ^65^, regression analysis was performed using R version R-4.0.3 ^66^, and visualization using ggplot2^67^.

## Results

### Characterization of the study population and infecting HIV-1 isolates

Overall, 375 plasma samples from 322 adult HIV-1 infected patients from three sampling years, 2001 (n=106), 2009 (n=210) and 2014 (n=59), were included in this analysis. Epidemiological, clinical, demographic, and virological characterization of the patients is described in **Table S1**. The median age of the patients was 34 years and most (n=242, 64.5%) were women. The main route of transmission was heterosexual contact (n=304, 81.1%). There were no significant differences related to age and sex between sampling years. The median plasma viral load (VL) at the time of sampling was significantly higher in 2001 relative to 2009 (4.2-fold higher) and 2014 (33.5-fold higher). The median number of CD4+ T cells in 2014 was 1.8-fold higher when compared to 2009 (*p*=0.0015). The significantly lower VL and higher CD4+ T cell number in 2014 is consistent with most patients being on cART which was not the case in 2001 and 2009.

Sequencing and phylogenetic analysis of the C2V3C3 Env region was completed successfully for 206 patients from 2001 (n=96/106, 90.6%) and 2009 (n=110/210, 52.4%). The following subtypes were identified: A1 (2001, n=33, 34.4%; 2009, n=32, 29.1%), A2 (2001, n=6, 6.3%; 2009, n=3, 2.7%), B (2001, n=2, 2.1%; 2009, n=2, 1.8%), C (2001, n=12, 12.5%; 2009, n=30, 27.3%), D (2001, n=2, 2.1%; 2009, n=8, 7.3%), F1 (2001, n=5, 5.2%; 2009, n=6, 5.5%), G (2001, n=8, 8.3%; 2009, n=11, 10%), H (2001, n=19, 19.8%; 2009, n=15, 13.6%), and J (2001, n=3, 3.1%; 2009, n=0, 0.0%). Untypable U strains were 4.2% (n=6) in 2001 and 2.7% (n=3) in 2009 (**Figure S1**). Subtype A prevailed in 2001 and 2009, but subtype C increased significantly (2.2-fold, *P=* 0.0095) in 2009. Out of the 176 isolates for which there were protease (PR) and C2V3C3 sequences available, 74 (42.0%) were non-recombinant and 102 (58.0%) were recombinant. Recombinant strains prevailed over pure subtypes in 2001 and 2009 (**Table S2**).

The genotypic analysis showed that most viruses were CCR5-tropic in 2001 (82.3%, N=79) and in 2009 (85.5%, N=94), without significant differences between sampling years (**Figure S2**). Unfortunately, we could not sequence the C2V3C3 region from most of the 2014 samples due to their low or undetectable viral load (**Table S1**). Moreover, the lack of plasma prevented further analysis in samples collected in 2001.

### Characterization of the antibody response

A total of 236 plasma samples, 178 from 2009 and 58 from 2014, were screened for neutralization breadth and potency against the 12 Env-pseudotyped indicator panel, amounting to 2832 plasma/virus combinations (**Figure S3**). In 2009, 80.9% (144/178) of Angolan patients effectively neutralized at least one virus from the indicator panel; this increased to 93.1% (54/58) in 2014 (**Figure 1**). Likewise, the mean percent neutralization (27.43%, 95%CI: 25.94, 28.92 in 2009 vs 60.52%, 95%CI: 57.37, 63.66 in 2014, p<0.0001) and the mean neutralization breadth (4.39, 95%CI: 3.83, 4.95 in 2009 vs 8.40, 95%CI: 7.32, 9.48 in 2014, p<0.0001) were higher in 2014 relative to 2009.

**Figure 1.**
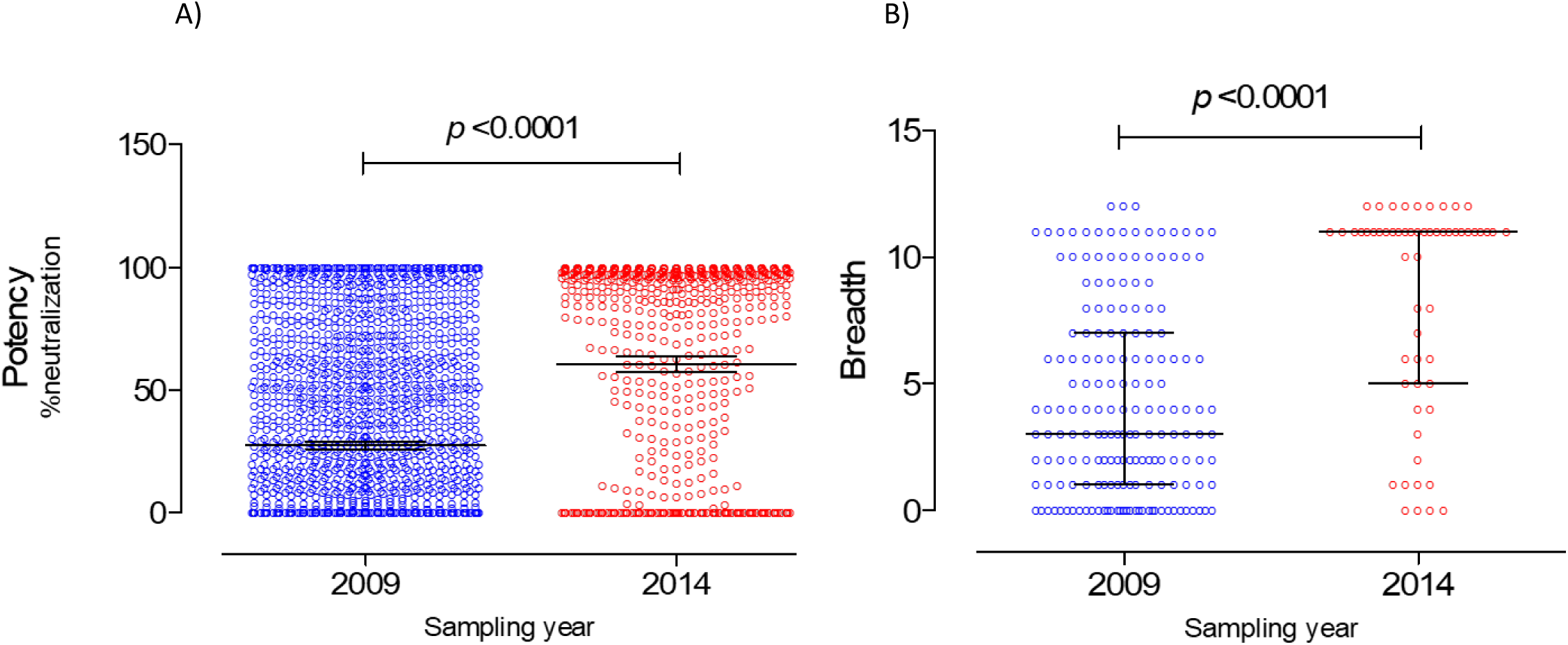
Neutralization potency and breadth per sampling year. A) Potency of neutralization (% neutralization at 1:40 plasma dilution) of samples collected in 2009 and 2014 as assessed against the 12-Env-pseudotyped virus indicator panel. Mean and 95% confidence intervals are shown. B) Neutralization breadth (number of Env-pseudotyped viruses neutralized >20%) in samples collected in 2009 and 2014. Median and interquartile range are shown P values were obtained using the Mann Whitney U test.

The percent neutralization for each plasma-virus combination was recorded as a breadth-potency matrix: ≥80% neutralization received a score of 3, 50% to <80% a score of 2, 20% to <50% a score of 1, and <20% received a score of 0. Plasma samples were then ranked by the sum of scores in order to reflect their potency and breadth ^2, 63^. Breadth, potency, and neutralization score were directly correlated as expected (**Figure S4**).

Mean NS was 11.71, 95% CI [10.22, 13.19] ranging from 0 to 36 and median was 7, IQR [2.0, 21.0]. Remarkably, approximately 30% (n= 68/236) of the patients developed antibody responses with the capacity to potently neutralize at least half the viruses from the panel (**Figure 2A**). Overall, considering both sampling years, 18.6% (n= 44/236) of study participants were elite neutralizers (NS ≥ 25), 10.2% (n= 24/236) were broad neutralizers (18 ≤ NS < 25), 27.1% (n= 64/236) were cross neutralizers (6 ≤ NS < 17), and 44.1% (n= 104/236) were weak neutralizers or did not neutralize any virus of the panel (**Figure 2B**).

**Figure 2.**
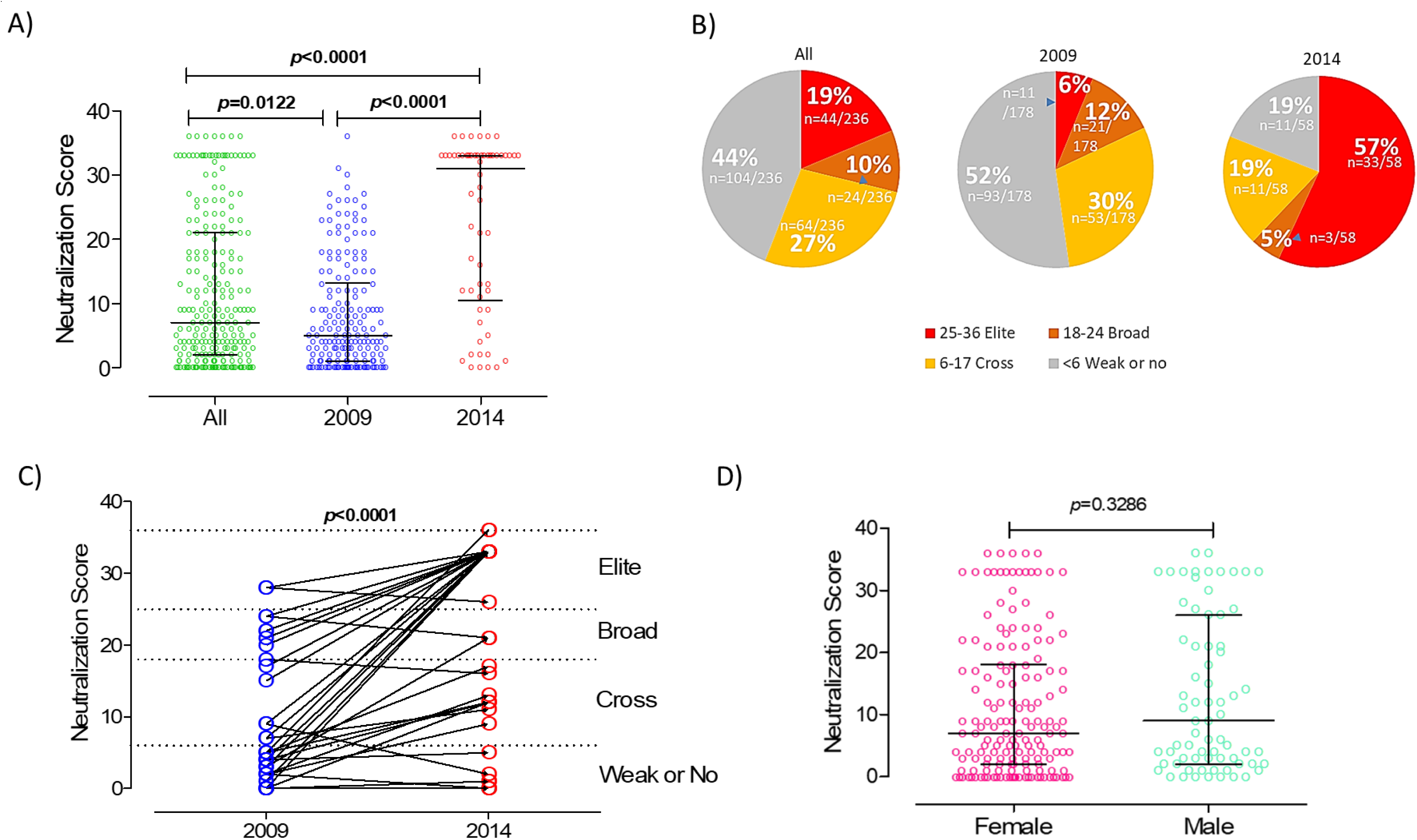
Neutralization score (NS) in HIV-1 infected patients from Angola as a function of year of sampling and sex. A) NS in 2009 is represented in blue and in 2014 in red; NS in all patients is in green. B) Angolan patients were categorized into 4 groups according to the NS as follows: no or weak neutralizers, <6 (grey); Cross neutralizers, 6-17 (yellow); Broad neutralizers, 18-24 (orange); Elite neutralizers, 25-36 (red). C) Neutralization score in matched samples collected in 2009 and 2014, showing the NS categories. D) NS in males and females. Median and interquartile range are shown. P values were obtained using the Mann Whitney U test.

### Correlates of the neutralizing response

The neutralizing antibody response has been previously associated with viral load, CD4+ T cell count, viral diversity and infection time ^2, 3^. We first analyzed the impact of sample collection time on the neutralizing antibody responses of the HIV infected Angolan patients. Strikingly, median NS was 6.2-fold higher in 2014 relative to 2009 (31.00, IQR [10.50, 33.00] vs 5.00, IQR [1.00, 13.25], p<0.0001) (**Figure 2A**). Consistent with this, the frequency of elite neutralizers was 9.5-fold higher in 2014 than in 2009 [57% (n=33/58) vs 6% (n= 11/178), *p*<0.0001], and weak or no neutralizers were 2.7-fold more frequent in 2009 than in 2014 [52% (n= 93/178) vs 19.0% (n= 11/58), *p*<0.0001] (**Figure 2B**). Broad neutralizers were 2.4-fold more frequent in 2009 than in 2014 [12% (n= 21/178) vs 5% (n= 3/58), *p*=0.2107] and a similar trend was observed for cross neutralizers [30% (n= 53/179) vs 19.0% (n= 11/58), *p*=0.1270]. We also analysed matching plasma pairs from 2009 and 2014 to determine the evolution of neutralizing antibody response as a function of infection time. In line with the previous results, neutralizing score increased in 2014 relative to 2009 in 31 out of the 38 matched plasma pairs analysed (81.6%). (**Figure 2C**). The NS was unrelated with the sex of the patients (**Figure 2D**).

The 50% neutralization titers (ID_50_) against the 12 Env-pseudotyped virus indicator panel were determined in a subset of plasma samples from 2009 (n=28) and 2014 (n=10) showing broad and elite neutralizing activity (**Figure 3A**). When comparing unmatched samples, neutralization titers were significantly higher in 2014 than in 2009 [median log_10_ ID_50_ in 2009= 1.903, IQ: 1.602-2.505 (n=336 plasma-virus pairs) vs median log_10_ ID_50_ in 2014= 2.204, IQ: 1.903-2.806 (n= 120 plasma-virus pairs), *p*=0.0013] (**Figure 3A/B**).

**Figure 3.**
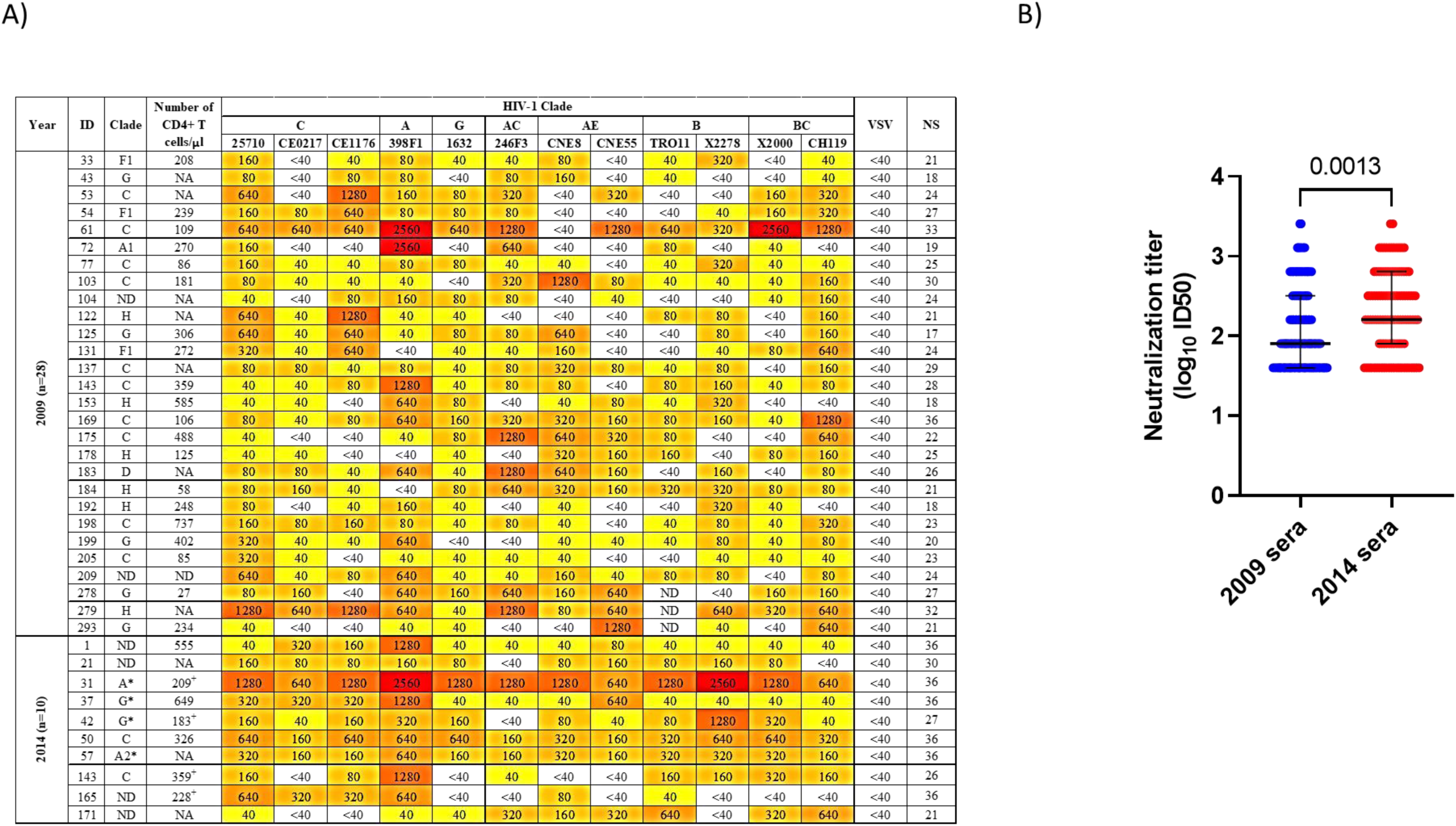
Antibody neutralization titers in a subset of unmatched plasma samples from elite and broad neutralizers from 2009 (n=28) and 2014 (n=10). A) Heatmap of the neutralization titers (ID50) and neutralization score against the 12 Env-pseudotyped virus indicator panel. ID50 values are color-coded, with darker colors implying higher ID50 values. *HIV subtype determined in the pol gene. ^+^ Number of CD4+ T cells determined in 2009; ND- not done due to lack of sample; NS-neutralization score; VSV-pseudotyped viruses (neutralization specificity control). B) Comparison of antibody neutralization titers in 2009 and 2014. Log_10_ ID50 values obtained by each patient sample against the 12 Env-pseudotyped virus indicator panel are plotted. Lines indicate the median with interquartile range. P value was obtained using the Mann Whitney U test.

Overall, these results suggest that duration of infection is an important correlate of the potency and breadth of the neutralizing antibody response in Angolan patients infected with HIV-1.

To analyse the impact of HIV-1 subtype on neutralization by Angolan samples we compared the neutralization score (NS) in patients infected with subtypes C (n=27) and A1 (n=26), the two prevailing subtypes in Angola, and in patients infected with the other subtypes and recombinant forms (n=56). Only samples collected in 2009 were included in this analysis due to the limited number of samples genotyped in 2014. NS varied significantly with infecting virus subtype (*p*=0.014), with subtype C leading to significantly higher NS than subtype A1 [median NS= 17.00, IQR (6.00, 25.00) vs 6.00 IQR (3.50, 15.00), *p*=0.0103] or other subtypes [median NS= 17.00, IQR (6.00, 25.00) vs 8.00, IQR (1.00, 17.75), *p*=0.0087] (**Figure 4A**). These results indicate that virus subtype is a major determinant of the neutralizing antibody response in our patients.

**Figure 4.**
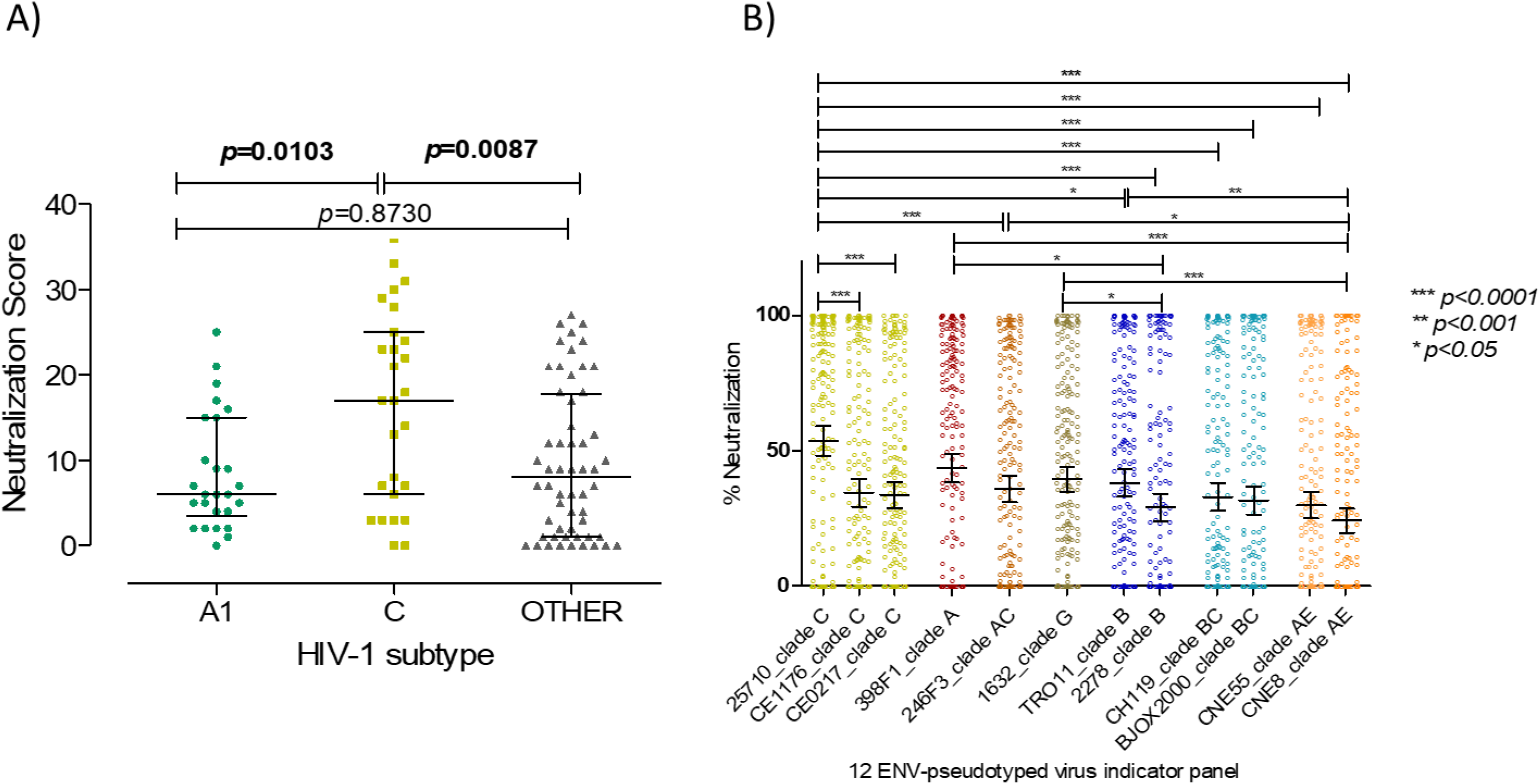
Impact of HIV-1 subtype on antibody neutralization. A) Neutralization score in patients infected with the two most common subtypes in Angola (year 2009), C (n=27) and A1 (n=26), and in patients infected with other subtypes and recombinant forms (n=56). The Kruskal-Wallis nonparametric test was used to analyse the difference in median NS for all subtypes (*p=*0.014). Dunns multiple comparison test was used to analyse differences in NS between subtypes. Median and interquartile range are shown. B) Percent neutralization of each of the 12 Env-pseudotyped virus indicator panel by the plasma samples (at 1:40 dilution) from the Angolan patients (N=236). Mean percent neutralization and 95% confidence interval bars against a given virus from the indicator panel are shown. Statistically significant differences are represented by the P values obtained with Dunns multiple comparison test. *** p<0.0001, ** p<0.001, *p<0.05.

The indicator virus panel used in the neutralization experiments contains three subtype C strains (25710, CE1176, and CE0217) that could be more closely related to the subtype C isolates from the Angolan patients and explain the higher NS observed in patients infected with subtype C viruses. To examine this issue, we compared the susceptibility of the reference panel isolates to neutralization and found a significant variation related to virus subtype (F**igure 4B**). The easiest viruses to neutralize were isolate 25710, a subtype C from India, and 398F1, a subtype A from Tanzania. On the other hand, viruses most resistant to neutralization were 2278, a subtype B from Spain and CNE8, a CRF01_AE from China. Interestingly, subtype C isolates CE1176 and CE0217 from Malawi were significantly more resistant to neutralization than 25710 suggesting a closer relationship of subtype C isolates from Angola to this Indian subtype C isolate and showing that subtype per is not the main determinant of susceptibility to antibody neutralization. To investigate the impact of the evolutionary distance between the HIV-1 Angolan isolates and the indicator virus panel on the neutralizing antibody responses, we aligned the C2V3C3 amino acid sequences from the Angolan isolates (year 2009) with those from the indicator virus panel. As expected, subtype C viruses from the patients were more closely related with subtype C viruses of the indicator panel relative to other subtypes (**Figure S5A**). There was a significant negative correlation of amino acid distance of the indicator panel to NS (Spearman r= -0.2319, *p* = 0.019) (**Figure S5B**). Hence, the closer the isolate from the indicator panel was to the patient’s C2V3C3 amino acid sequence, the easier it was neutralized. On average, clade C reference strain 25710 from the indicator panel was the closest indicator virus panel member to the Angolan isolates and, not surprisingly, it was the easiest virus to neutralize. At the other end, clade B reference strain 2278 was the furthest away from the C2V3C3 Angolan sequences and was the most difficult virus to neutralize along with the CRF01_AE virus (CNE8). Nevertheless, many patients infected with all subtypes developed potent bNAb responses despite the high genetic distance to the viruses of the indicator panel, indicating that other factors besides the relatedness with the indicator panel contribute to the potency of the neutralizing response.

Drug naïve patients (year 2009) with ≤200 CD4+ T cells/μl at study entry had significantly higher NS values than patients with >200 CD4+ T cells/μl [median NS in patients ≤200 CD4+ T cell counts was 7.00 (IQR, 3.50, 21.00) vs 4.00 (1.00, 12.00) in patients with > 200 CD4+ T cell counts, p = 0.0193] (**Figure 5A**). Moreover, NS values were inversely associated with CD4+ T cell counts (Spearman r=-0.3043, *p*=0.0005) **(Figure 5B)** and directly associated with age (Spearman r=0.1644, p =0.0302) in these patients (**Figure 5C)**. These results suggest that elicitation of high levels of broadly neutralizing antibodies in these patients is directly correlated with prolonged antigenic stimulation^68^.

**Figure 5.**
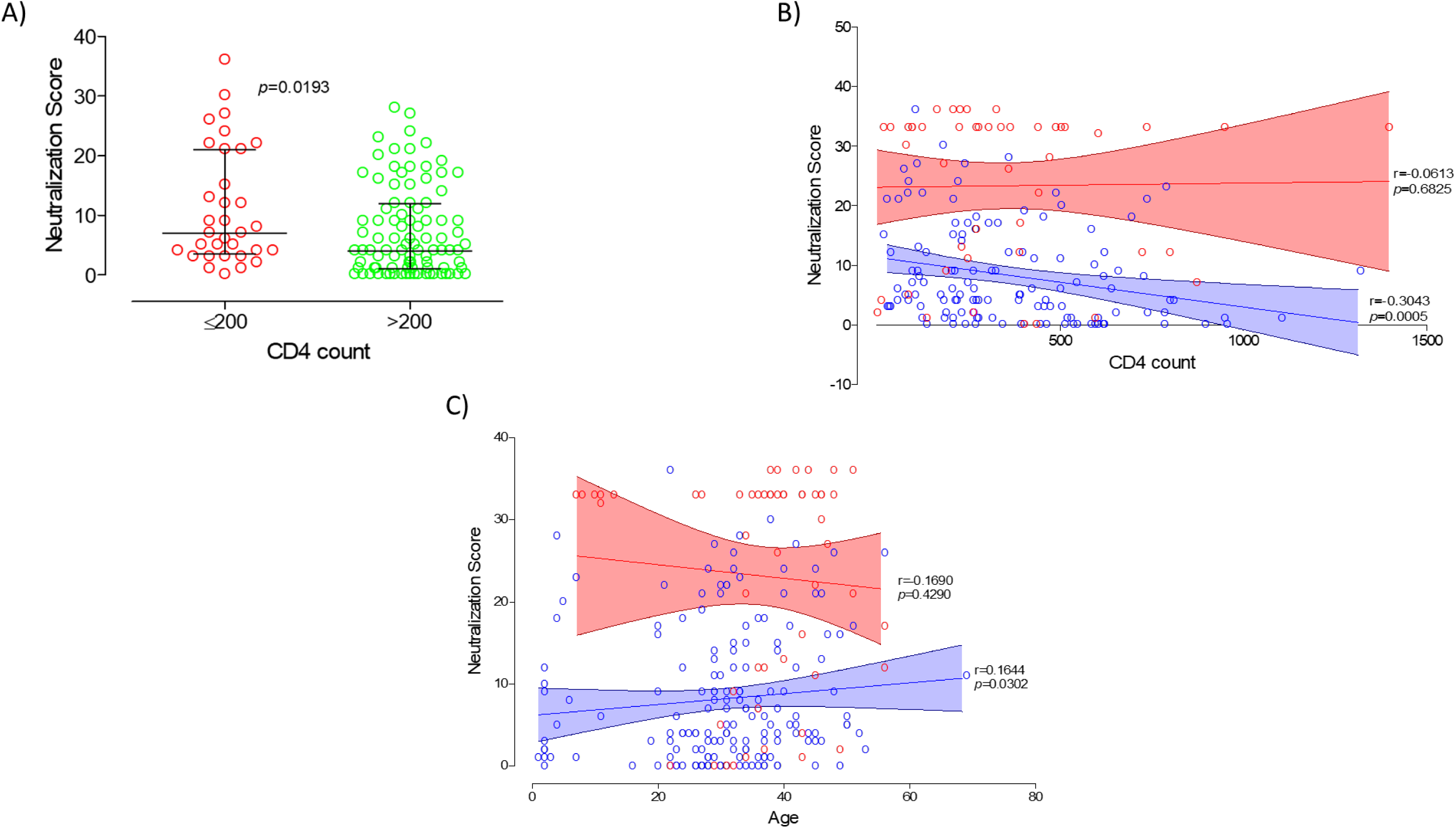
Correlation between neutralization score, CD4+ T cell counts and patient’s age. A) Neutralization score differences between 2009 patient’s with ≤ 200 CD4+ T cell counts at study entry and patients with >200 CD4 T cell counts. Median and interquartile range are shown. P values were obtained using the Mann Whitney U test; B) Correlation of neutralization score with CD4+ T cell counts in 2009 and 2014; C) Correlation of neutralization score with patient’s age in 2009 and 2014. Samples collected in 2009 are shown in blue and samples collected in 2014 in red. Linear trend is shown with mean and 95% CI bands; Spearman r and P values are indicated.

### Epitope specificities of the plasma neutralizing antibodies

In the same subset of 38 plasma samples (n=28 from 2009 and n=10 from 2014) from broad and elite neutralizers, the epitope specificities were mapped using a computational clustering tool based on the epitope specificities of a panel of human bNAbs ^64^. Six (15.8%) samples did not cluster with any of the bNAbs. Thirty-two (84.2%) samples clustered with one of the bNAbs. Of these most samples (68.8%, 22/32) clustered with PGT128 and 2G12, two bnAbs that target the V3 glycan supersite with important contact residues in V3 and V4 (**Figure 6**) ^69, 70^. Five (15.6%) samples clustered with bnAb 4E10 that targets the gp41 membrane-proximal external region (MPER) ^71^. Four (12.5%) samples clustered with VRC01 and VRC-CH31 bnAbs that target the CD4 binding site. Finally, one (3.1%) sample clustered with PG16 and PG9 that target the V1V2 glycans. These results indicate that the V3 glycan supersite is the dominant broadly neutralizing epitope in Angolan patients.

**Figure 6.**
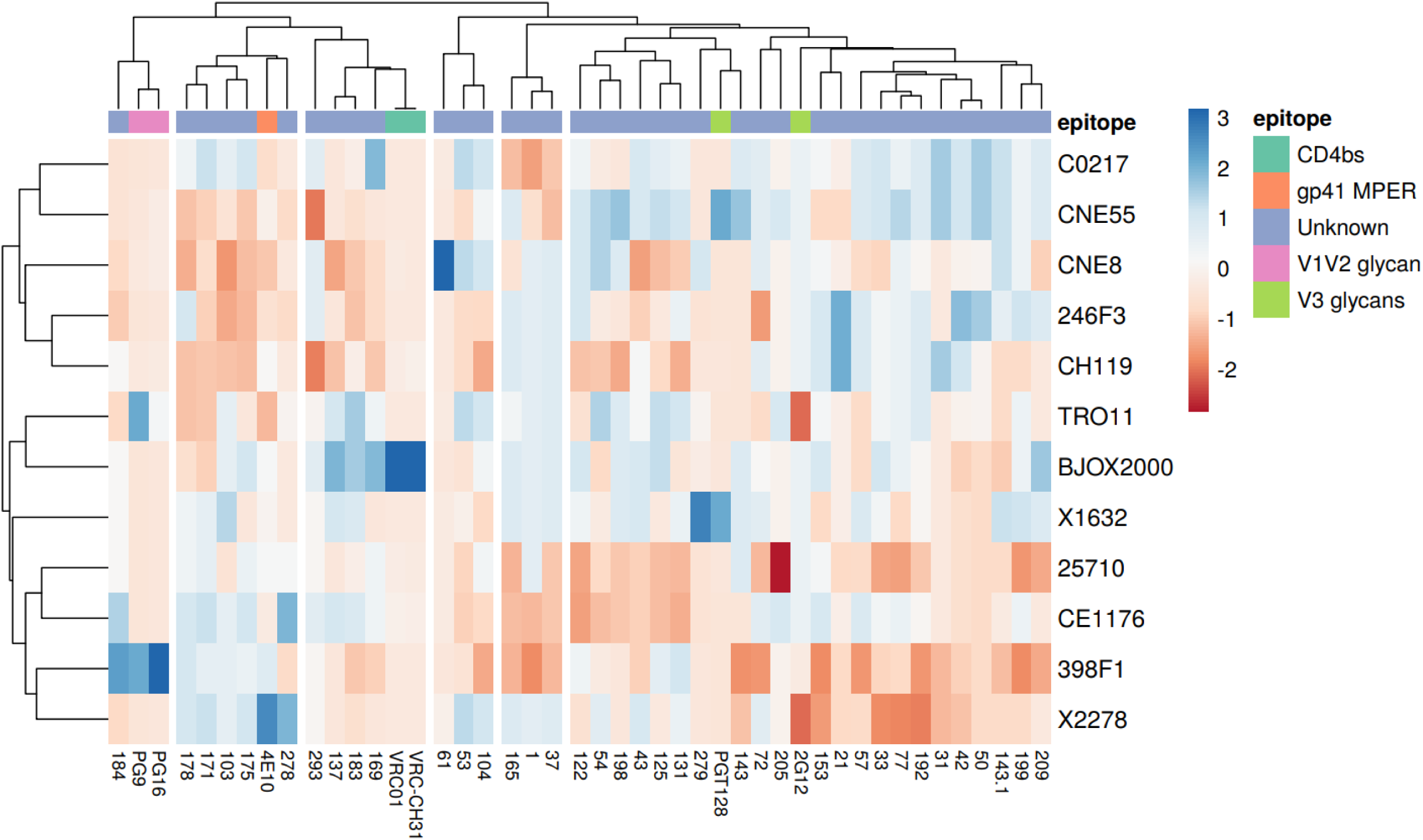
**Cluster analysis and heatmap of the predicted epitope specificity in the top neutralizing patients from Angola.** In the top of the columns, known bnAb epitopes are coloured according to the respective epitope specificities as shown by the legend. The identification of the plasma samples and bnAbs is shown in the bottom of the columns. Cluster analysis for both rows and columns were computed according to the Pearson correlation ^64^. Blue colours in the heatmap represent lower neutralization activity and red colours higher neutralization activity. Each column represents the neutralization values of a given plasma sample or a bnAb of known specificities against the 12 Env-pseudotyped virus panel whose names are indicated to the left.

### Neutralization score is directly related with titer of C2V3C3-binding antibodies in all subtypes

The antibody binding reactivity against a panel of recombinant polypeptides comprising the C2, V3, and C3 envelope regions of subtypes B, C, G, H, J, and CRF02_AG was characterized in a subset of samples from 2009 (n=48) and 2014 (n=16) with known antibody neutralization profile. All but the B polypeptide were derived from Angolan isolates. All but six samples from five patients reacted with all C2V3C3 polypeptides demonstrating the high antigenicity of this envelope region (**Figure S6**). In 2009, patients had significantly higher median antibody binding titers against subtype C than against subtypes G (*p*=0.0007), H (*p*=0.0282), J (*p*=0.0052), and CRF02_AG (*p*=0.0149). Of note, median antibody binding titers were always higher in 2014 relative to 2009 regardless of the C2V3C3 polypeptide subtype but this was not significant except for CRF02_AG.

The higher antibody reactivity against subtype C antigen could be related with the higher neutralizing responses observed in subtype C infected patients. We therefore investigated possible associations between neutralization score, C2V3C3 antibody binding titer and subtype. Remarkably, C2V3C3 antibody binding titer was positively associated with NS values, i.e., patients with higher antibody binding titers to C2V3C3 polypeptides had higher neutralizing antibody responses (F**igure 7**). This was significant for all subtypes of the C2V3C3 recombinant polypeptides tested confirming this epitope as an important neutralizing domain in these patients independent of subtype.

**Figure 7.**
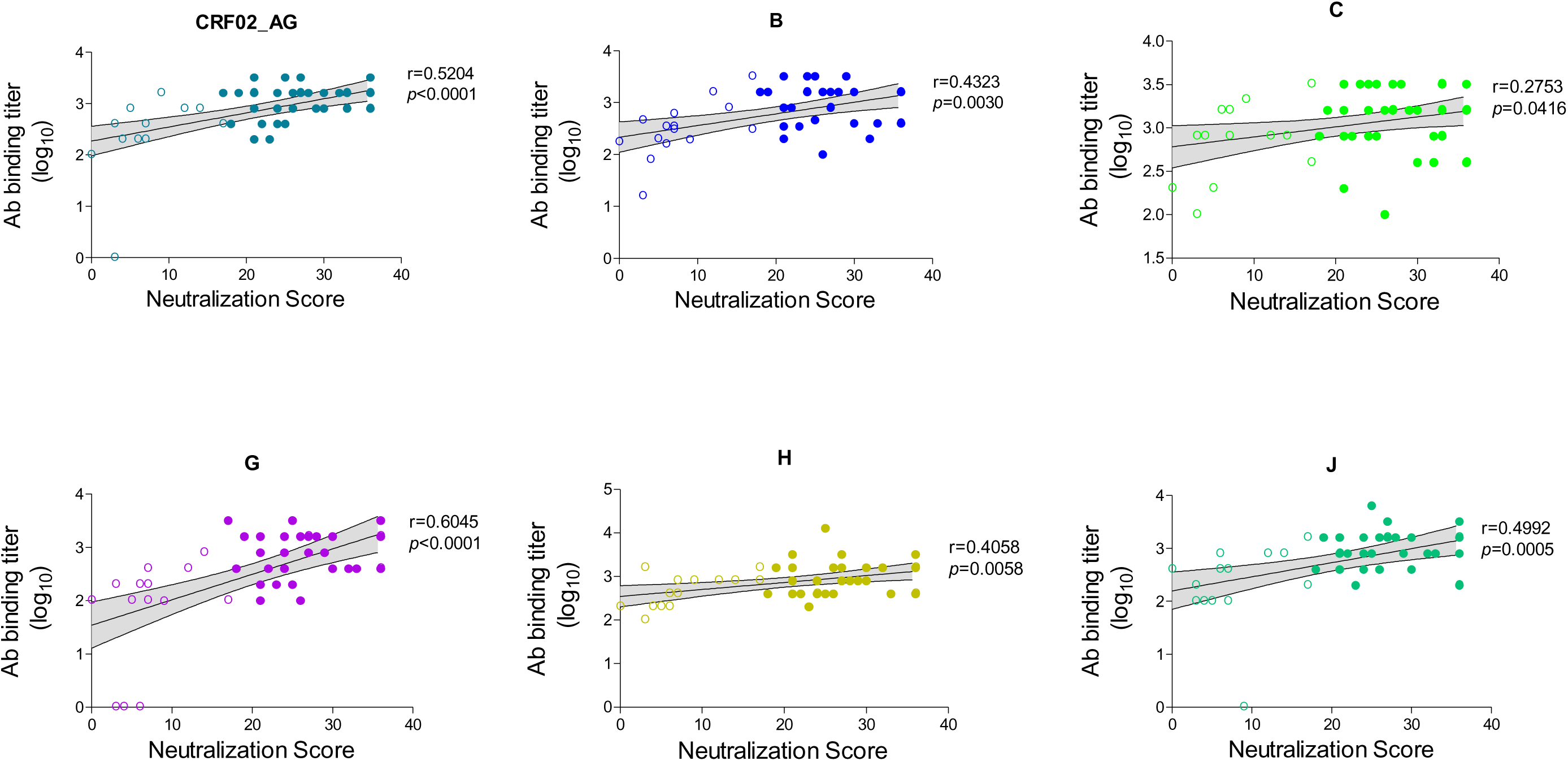
Association between antibody binding titer to C2V3C3 recombinant polypeptides of different subtypes and neutralization score (year 2009). Filled symbols are the antibody binding titers of the broad/elite neutralizers to a given C2V3C3 subtype. Unfilled symbols are the antibody binding titers of the no/weak and cross neutralizers to a given C2V3C3 subtype. Associations were assessed by Spearman analyses. P-values and Spearman r values are indicated. Linear trend is shown with mean and 95% CI bands.

### Impact of the neutralizing antibodies in the diversity and evolution of C2V3C3

Neutralizing antibodies targeting the C2, V3 and C3 envelope regions are common in HIV-1 infected individuals ^3^ and escape from these antibodies leads to higher diversity in these regions as well as to higher positive selection and convergent evolution ^72, 73, 74^. To investigate the impact of the neutralizing antibodies in the diversity and evolution of the envelope glycoproteins of the viruses infecting our patients, we analysed amino acid entropy and the sites under selective pressure in the C2, V3 and C3 regions in the different neutralization categories for samples collected in 2009. Considering the three regions together, mean overall entropy values were similar in all neutralization categories: weak/no neutralizers= 0.5727 [95% confidence interval (CI): 0.4753, 0.6241]; cross neutralizers= 0.5931 (0.5012, 0.6849); broad neutralizers= 0.5381 (0.4472, 0.6291); and elite neutralizers= 0.4106 (0.3313, 0.4899). Regardless of neutralization category, the region with higher mean entropy was C3 [0.8528 (0.7556, 0.9500)] followed by V3 [0.4659 (0.3903, 0.5414)] and C2 [0.3635 (0.3092, 0.4178)] (*p*<0.0001). We then plotted Shannon’s entropy differences in C2V3C3 between no/weak neutralizers and cross, broad and elite neutralizers. This analysis revealed that viruses from elite neutralizers were far less variable than viruses from weak/no neutralizers as seen by the number of amino acids with positive entropy differences relative to amino acids with negative entropy differences (37 vs 16 sites, p=0.0023) (**Figure 8**). On the other hand, viruses from broad and cross neutralizers did not vary significantly from viruses from no/weak neutralizers. Relative to no/weak neutralizers, the most variable amino acid residues in the broad and elite neutralizers were found in V3 and/or C3 (broad neutralizers: 1 site in C2 vs 7 sites in C3, p<0.0001; elite neutralizers: 3 sites in C2 vs 13 sites in V3C3, p<0.0001), two regions that contain broadly neutralizing epitopes (**Figure 8**).

**Figure 8.**
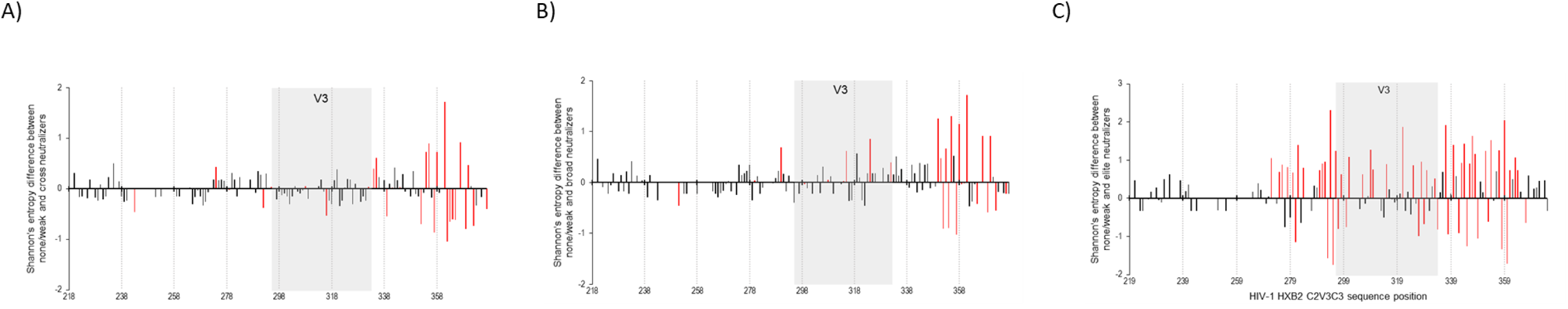
Amino acid entropy difference in the C2V3C3 region between neutralization categories. A) Shannon’s entropy difference between No/Weak and Cross neutralizers. B) Shannon’s entropy difference between No/Weak and Broad neutralizers. C) Shannon’s entropy difference between No/Weak and Elite neutralizers. Sites with significant entropy difference (*p* ≤0.05) are shown in red. Gray boxes delimitate the V3 region. Numbers in the x axes indicate the amino acid position in HIV-1 HXB2.

Diversifying selection in C2V3C3 varied according to the different neutralization categories (*p*<0.001) (**Table S3**). Considering only sites that were selected by at least two methods, weak/no neutralizers had a total of 9 positively selected sites, cross neutralizers 6, broad neutralizers 3 and elite neutralizers 1. Regardless of neutralization category most sites under selective pressure were present in the C3 region.

The mean number of N-glycosylation sites in C2V3C3 was similar in all neutralization categories [Non-neutralizers: 9.2 (range: 8-12); cross-neutralizers: 9.2 (range: 7-11), broad-neutralizers: 9.4 (range: 7-11); elite-neutralizers: 9.8 (range: 8-11)] (**Table S4**). C3 had more potential N-glycosylation sites than C2 or V3 but sites in V3 and C2 were more conserved. For example, sites 241, 262, 276 and 289 in C3 were present in ≥70% of strains and site 301 in the V3 crown was present in all but two strains (97%). In C3, site 332, which together with site 301 in V3 and other elements in V1, V3 and V4 is part of the V3-glycan supersite ^75, 76^, was highly conserved (70%) in all neutralization categories.

## Discussion

HIV-1 was introduced in Angola from Kinshasa, the capital city of the Democratic Republic of Congo (DRC), likely in 1910-1940 making the Angolan epidemic the second oldest in the world^48^. Like in the DRC, the Angolan epidemic has been driven by all subtypes but B, untypable and highly divergent strains, and multiple CRFs and URFs ^44, 45, 46, 47^. In this study we confirmed the extremely high diversity and evolving complexity of HIV-1 strains present in Angola. Subtypes A and C dominated over other subtypes but all other Env subtypes were present along with untypable basal strains and recombinant strains that prevailed over pure subtypes. The remarkable diversity and evolution of HIV-1 in Angola is driven by the increasing number of new infections ^77^, the limited access to antiretroviral therapy, and high levels of drug resistance ^46, 78^. The high diversity and rapid evolution of HIV-1 in this country can pose a serious challenge to vaccination and other preventive efforts.

At the individual level, the long-term B cell stimulation by this highly diversified ensemble of viruses may have promoted the development of exceptional neutralization breadth ^2, 3, 75, 79, 80, 81, 82, 83^. We found that the majority (56%) of the patients in our cohort developed cross, broad, or elite neutralizing responses. These results far exceed those from previous cohort studies in sub-Saharan Africa ^3, 63, 79, 84, 85^. For example, Beirnaert *et al*. found 10.6% broad neutralizers in Cameroon^85^ and Landais *et al*. found about 15% broad neutralizers in a cohort of HIV-1 infected patients from Eastern and South Africa^3^. When compared to cohort studies from other geographies where subtype B dominates, the frequency of patients with bNAb responses reported in our study was also much higher^2, 5, 86^. For example, Rusert *et al*. ^2^ in Switzerland found that most patients (79.1%) showed weak or no neutralization breadth, which compares to 44% in our cohort, and that only 1.3% were elite neutralizers which compares to 19% in our cohort. This divergence may be related with many factors besides the diversity of infecting viruses, such as the HLA genotype and ethnicity of the patients, viral load, CD4+ T cell counts, and duration of infection ^2, 3, 79, 80, 87^.

In agreement with other studies, neutralization score was inversely correlated with CD4+ T cell counts in 2009 when the patients were naïve to ART and had high viral load^2, 3, 75, 80^. This is generally associated with high envelope stimulation of B cells and inevitably leads to B-cell exhaustion in chronic viraemic HIV-1 infection ^88^. Remarkably, however, the frequency of elite neutralizers and the mean neutralization score in matched and unmatched patients increased significantly in 2014, when patients were already undergoing ART, relative to 2009. The boost in the quality of the neutralizing response in these patients suggest good restoration of the B cell compartment with ART which is uncommon in chronic HIV-1 infection ^88, 89, 90^. Moreover, the moderate level of plasma viremia (median 11,660 HIV-1 RNA copies /ml, IQR, 380-30,060) found in these patients may have provided the low-level antigenic stimulation needed for the full maturation of memory B cells and bNAb production ^31, 88, 91^. This has precedent in HIV-2 infection where most patients are infected for long periods and produce potent and broadly neutralizing responses in a setting of low plasma viremia^36, 92, 93^. In this model, B cell exposure to low-level envelope antigens, likely in lymphoid tissues, during prolonged infection periods leads to the generation of highly specific envelope C2V3C3-specific antibodies as well as broad and potent neutralizing antibodies ^94^.

Viral type and subtype as well as the nature of the epitope target on the viral envelope impact the antibody maturation process as seen by the frequency of elicitation ^61, 93^ and epitope specificity of bnAbs ^2, 50, 76^. Differences in envelope structure and epitope exposure, length of variable loops, type of V3 motifs, N-glycosylation patterns, and conservation of key sites have helped to explain why certain HIV-1 subtypes like subtype C are better at promoting the elicitation of neutralizing antibodies ^2, 3, 50, 61, 76, 84, 95^. In line with these studies, we found that infection with subtype C viruses was associated with enhanced neutralization breadth and potency. In general, subtype C infected individuals have shown a bias to V2-glycan directed antibody responses, and subtype C envelope from transmitted viruses have been less prone to neutralization by V3-directed antibodies due to the absence of the N332-glycan in the C3 region ^2, 3, 76, 84, 96, 97, 98^. This was not the case in our study as most top neutralizers had V3-directed antibodies that were able to neutralize the subtype C isolates from the virus panel, and the N301 and N332 glycans defining the V3-glycan supersite were highly conserved in the patient’s isolates. Supporting the major role of the V3 and C3 envelope regions in the development of bNAbs in our cohort, we found a strong direct correlation between the titer of antibodies binding to C2V3C3 envelope polypeptides from all subtypes and neutralization score. Nonetheless, antibodies specific for the V2 apex, the CD4 binding site, the gp41 MPER and/or unknown epitopes were also found in some patients revealing the complexity of the neutralizing antibody responses in these patients.

We also looked at the variability of patient’s sequences in the envelope C2V3C3 region to assess the impact of escape from neutralizing antibodies on viral evolution and diversity. V3 and C3 were the most variable regions which is consistent with the dominant role of neutralizing antibodies targeting these regions in these patients ^3^. V3 bNAb recognition sites and sites associated with resistance to neutralization such as N295 ^3, 75, 80^ were under positive selection in broad and elite neutralizers. However, elite neutralizers exhibited far less variability and lower number of sites under selective pressure in V3 and C3 relative to weak or low neutralizers. The convergence of the viral swarm to a few resistant strains provides convincing evidence for the crucial role of V3- and C3-directed bNAbs in controlling HIV-1 replication and diversification in these patients ^35, 61, 84, 99^.

In conclusion, an exceptionally high number of Angolan patients infected with HIV-1 elicits broad and elite neutralizing antibodies mostly targeting the V3-glycan supersite. This is associated with long-term and low-level V3- and C3- antigenic stimulation by the highly diverse isolates circulating in this country especially subtype C. These results have direct implications for the design of a new generation of HIV-1 vaccines.

## Data Availability

All data produced in the present study are available upon reasonable request to the authors

## Acknowledgments

We greatly acknowledge the contribution and efforts of the staff and patients from the Hospital da Divina Providência in Luanda for this study. TZM-bl cells were obtained through the NIH AIDS Reagent Program, Division of AIDS, NIAID, NIH. This work was supported by Fundação para a Ciência e a Tecnologia (FCT), Portugal, under project grants UIDB/04138/2020 and UIDP/04138/2020. This study was in part supported by the NIH/NIAID under grant R01AI087520. Francisco Martin was supported by FCT under PhD grant number SFRH/BD/87488/2012. CP is funded by FCT under a contract-program as defined by DL No. 57/2016 and Law No. 57/2017.

## Authors’ contributions

Francisco Martin^1^, José Maria Marcelino^1,2^, Claudia Palladino^1^, Inês Bártolo^1^, Susana Tracana^1^, Inês Moranguinho^1^, Paloma Gonçalves^1^, Rita Mateus^1^, Rita Calado^1^, Pedro Borrego^1^, Thomas Leitner^3^, Sofia Clemente^4^, Nuno Taveira^1,2#^

NT and SC: conceived and designed the study. SC: provided the patients’ data and specimens. FM, JMM, IB, ST, IM, PG, RM, RC, PB, and TL: performed the experiments. FM, JMM, CP, TL and NT: analyzed the data. FM, TL, and NT performed the statistical analysis. FM, TL and NT drafted the manuscript and discussed the final version. All authors read and approved the final manuscript.

## References

1. Horwitz JA, et al. Non-neutralizing Antibodies Alter the Course of HIV-1 Infection In Vivo. Cell 170, 637–648 e610 (2017).

2. Rusert P, et al. Determinants of HIV-1 broadly neutralizing antibody induction. Nat Med 22, 1260–1267 (2016).

3. Landais E, et al. Broadly Neutralizing Antibody Responses in a Large Longitudinal Sub-Saharan HIV Primary Infection Cohort. PLoS Pathog 12, e1005369 (2016).

4. Hu X, et al. Profiling the neutralizing antibody response in chronically HIV-1 CRF07_BC- infected intravenous drug users naive to antiretroviral therapy. Sci Rep 7, 46308 (2017).

5. Hraber P, et al. Impact of clade, geography, and age of the epidemic on HIV-1 neutralization by antibodies. J Virol 88, 12623–12643 (2014).

6. Tomaras GD, et al. Polyclonal B cell responses to conserved neutralization epitopes in a subset of HIV-1-infected individuals. J Virol 85, 11502–11519 (2011).

7. Simek MD, et al. Human immunodeficiency virus type 1 elite neutralizers: individuals with broad and potent neutralizing activity identified by using a high-throughput neutralization assay together with an analytical selection algorithm. J Virol 83, 7337–7348 (2009).

8. Stephenson KE, Wagh K, Korber B, Barouch DH. Vaccines and Broadly Neutralizing Antibodies for HIV-1 Prevention. Annu Rev Immunol 38, 673–703 (2020).

9. Walker LM, et al. Broad and potent neutralizing antibodies from an African donor reveal a new HIV-1 vaccine target. Science 326, 285–289 (2009).

10. Caskey M, et al. Viraemia suppressed in HIV-1-infected humans by broadly neutralizing antibody 3BNC117. Nature 522, 487–491 (2015).

11. Caskey M, et al. Corrigendum: Viraemia suppressed in HIV-1-infected humans by broadly neutralizing antibody 3BNC117. Nature, (2016).

12. Caskey M, et al. Antibody 10-1074 suppresses viremia in HIV-1-infected individuals. Nat Med 23, 185–191 (2017).

13. Schoofs T, et al. HIV-1 therapy with monoclonal antibody 3BNC117 elicits host immune responses against HIV-1. Science 352, 997–1001 (2016).

14. Scheid JF, et al. HIV-1 antibody 3BNC117 suppresses viral rebound in humans during treatment interruption. Nature 535, 556–560 (2016).

15. Nishimura Y, Martin MA. Of Mice, Macaques, and Men: Broadly Neutralizing Antibody Immunotherapy for HIV-1. Cell Host Microbe 22, 207–216 (2017).

16. Corey L, et al. Two Randomized Trials of Neutralizing Antibodies to Prevent HIV-1 Acquisition. N Engl J Med 384, 1003–1014 (2021).

17. Spencer DA, Shapiro MB, Haigwood NL, Hessell AJ. Advancing HIV Broadly Neutralizing Antibodies: From Discovery to the Clinic. Front Public Health 9, 690017 (2021).

18. Burton DR, Mascola JR. Antibody responses to envelope glycoproteins in HIV-1 infection. Nat Immunol 16, 571–576 (2015).

19. Gray GE, et al. Vaccine Efficacy of ALVAC-HIV and Bivalent Subtype C gp120-MF59 in Adults. N Engl J Med 384, 1089–1100 (2021).

20. Sanders RW, et al. A next-generation cleaved, soluble HIV-1 Env trimer, BG505 SOSIP.664 gp140, expresses multiple epitopes for broadly neutralizing but not non-neutralizing antibodies. PLoS Pathog 9, e1003618 (2013).

21. Julien JP, et al. Design and structure of two HIV-1 clade C SOSIP.664 trimers that increase the arsenal of native-like Env immunogens. Proc Natl Acad Sci U S A 112, 11947–11952 (2015).

22. Sanders RW, et al. HIV-1 VACCINES. HIV-1 neutralizing antibodies induced by native-like envelope trimers. Science 349, aac4223 (2015).

23. Pugach P, et al. A native-like SOSIP.664 trimer based on an HIV-1 subtype B env gene. J Virol 89, 3380–3395 (2015).

24. Zhang P, et al. A multiclade env-gag VLP mRNA vaccine elicits tier-2 HIV-1-neutralizing antibodies and reduces the risk of heterologous SHIV infection in macaques. Nat Med 27, 2234–2245 (2021).

25. Julg B, Barouch DH. Neutralizing antibodies for HIV-1 prevention. Curr Opin HIV AIDS 14, 318–324 (2019).

26. Cohen YZ, Caskey M. Broadly neutralizing antibodies for treatment and prevention of HIV-1 infection. Curr Opin HIV AIDS 13, 366–373 (2018).

27. McGuire AT, Glenn JA, Lippy A, Stamatatos L. Diverse recombinant HIV-1 Envs fail to activate B cells expressing the germline B cell receptors of the broadly neutralizing anti-HIV-1 antibodies PG9 and 447-52D. J Virol 88, 2645–2657 (2014).

28. McGuire AT, et al. Specifically modified Env immunogens activate B-cell precursors of broadly neutralizing HIV-1 antibodies in transgenic mice. Nat Commun 7, 10618 (2016).

29. Hu Y, et al. Virus Evolution and Neutralization Sensitivity in an HIV-1 Subtype B’ Infected Plasma Donor with Broadly Neutralizing Activity. Vaccines (Basel) 9, (2021).

30. Kwong PD, Mascola JR. HIV-1 Vaccines Based on Antibody Identification, B Cell Ontogeny, and Epitope Structure. Immunity 48, 855–871 (2018).

31. Cizmeci D, et al. Distinct clonal evolution of B-cells in HIV controllers with neutralizing antibody breadth. Elife 10, (2021).

32. Bonsignori M, et al. Staged induction of HIV-1 glycan-dependent broadly neutralizing antibodies. Sci Transl Med 9, (2017).

33. Simonich CA, et al. HIV-1 Neutralizing Antibodies with Limited Hypermutation from an Infant. Cell 166, 77–87 (2016).

34. Anthony C, et al. Cooperation between Strain-Specific and Broadly Neutralizing Responses Limited Viral Escape and Prolonged the Exposure of the Broadly Neutralizing Epitope. J Virol 91, (2017).

35. Martinez DR, et al. Maternal Binding and Neutralizing IgG Responses Targeting the C-Terminal Region of the V3 Loop Are Predictive of Reduced Peripartum HIV-1 Transmission Risk. J Virol 91, (2017).

36. Marcelino JM, et al. Resistance to antibody neutralization in HIV-2 infection occurs in late stage disease and is associated with X4 tropism. AIDS 26, 2275–2284 (2012).

37. Rocha C, et al. Evolution of the human immunodeficiency virus type 2 envelope in the first years of infection is associated with the dynamics of the neutralizing antibody response. Retrovirology 10, 110 (2013).

38. Serra PA, Taveira N, Guedes RC. Computational Modulation of the V3 Region of Glycoprotein gp125 of HIV-2. Int J Mol Sci 22, (2021).

39. Gray ES, et al. Antibody specificities associated with neutralization breadth in plasma from human immunodeficiency virus type 1 subtype C-infected blood donors. J Virol 83, 8925–8937 (2009).

40. Kostrikis LG, Cao Y, Ngai H, Moore JP, Ho DD. Quantitative analysis of serum neutralization of human immunodeficiency virus type 1 from subtypes A, B, C, D, E, F, and I: lack of direct correlation between neutralization serotypes and genetic subtypes and evidence for prevalent serum-dependent infectivity enhancement. J Virol 70, 445–458 (1996).

41. Moore JP, Cao Y, Leu J, Qin L, Korber B, Ho DD. Inter- and intraclade neutralization of human immunodeficiency virus type 1: genetic clades do not correspond to neutralization serotypes but partially correspond to gp120 antigenic serotypes. J Virol 70, 427–444 (1996).

42. deCamp A, et al. Global panel of HIV-1 Env reference strains for standardized assessments of vaccine-elicited neutralizing antibodies. J Virol 88, 2489–2507 (2014).

43. Marcelino R, et al. Antibody response against selected epitopes in the HIV-1 envelope gp41 ectodomain contributes to reduce viral burden in HIV-1 infected patients. Sci Rep 11, 8993 (2021).

44. Bartolo I, Calado R, Borrego P, Leitner T, Taveira N. Rare HIV-1 subtype J genomes and a new H/U/CRF02_AG recombinant genome suggests an ancient origin of HIV-1 in Angola. AIDS Res Hum Retroviruses, (2016).

45. Abecasis A, et al. HIV-1 genetic variants circulation in the North of Angola. Infect Genet Evol 5, 231–237 (2005).

46. Bartolo I, et al. HIV-1 diversity, transmission dynamics and primary drug resistance in Angola. PLoS One 9, e113626 (2014).

47. Bartolo I, et al. Highly divergent subtypes and new recombinant forms prevail in the HIV/AIDS epidemic in Angola: new insights into the origins of the AIDS pandemic. Infect Genet Evol 9, 672–682 (2009).

48. Pineda-Pena AC, et al. On the contribution of Angola to the initial spread of HIV-1. Infect Genet Evol 46, 219–222 (2016).

49. Faria NR, et al. HIV epidemiology. The early spread and epidemic ignition of HIV-1 in human populations. Science 346, 56–61 (2014).

50. Stefic K, Bouvin-Pley M, Braibant M, Barin F. Impact of HIV-1 Diversity on Its Sensitivity to Neutralization. Vaccines (Basel) 7, (2019).

51. Platt EJ, Wehrly K, Kuhmann SE, Chesebro B, Kabat D. Effects of CCR5 and CD4 cell surface concentrations on infections by macrophagetropic isolates of human immunodeficiency virus type 1. J Virol 72, 2855–2864 (1998).

52. Bartolo I, et al. HIV-1 genetic diversity and transmitted drug resistance in health care settings in Maputo, Mozambique. J Acquir Immune Defic Syndr 51, 323–331 (2009).

53. Tamura K, Stecher G, Peterson D, Filipski A, Kumar S. MEGA6: Molecular Evolutionary Genetics Analysis version 6.0. Mol Biol Evol 30, 2725–2729 (2013).

54. Posada D, Crandall KA. MODELTEST: testing the model of DNA substitution. Bioinformatics 14, 817–818 (1998).

55. Guindon S, Dufayard JF, Lefort V, Anisimova M, Hordijk W, Gascuel O. New algorithms and methods to estimate maximum-likelihood phylogenies: assessing the performance of PhyML 3.0. Syst Biol 59, 307–321 (2010).

56. Lengauer T, Sander O, Sierra S, Thielen A, Kaiser R. Bioinformatics prediction of HIV coreceptor usage. Nat Biotechnol 25, 1407–1410 (2007).

57. Vandekerckhove LP, et al. European guidelines on the clinical management of HIV-1 tropism testing. Lancet Infect Dis 11, 394–407 (2011).

58. Weaver S, Shank SD, Spielman SJ, Li M, Muse SV, Kosakovsky Pond SL. Datamonkey 2.0: A Modern Web Application for Characterizing Selective and Other Evolutionary Processes. Mol Biol Evol 35, 773–777 (2018).

59. Kosakovsky Pond SL, Frost SD. Not so different after all: a comparison of methods for detecting amino acid sites under selection. Mol Biol Evol 22, 1208–1222 (2005).

60. Zhang M, et al. Tracking global patterns of N-linked glycosylation site variation in highly variable viral glycoproteins: HIV, SIV, and HCV envelopes and influenza hemagglutinin. Glycobiology 14, 1229–1246 (2004).

61. Calado R, et al. A Prime-Boost Immunization Strategy with Vaccinia Virus Expressing Novel gp120 Envelope Glycoprotein from a CRF02_AG Isolate Elicits Cross-Clade Tier 2 HIV-1 Neutralizing Antibodies. Vaccines (Basel) 8, (2020).

62. Mascola JR, et al. Recommendations for the design and use of standard virus panels to assess neutralizing antibody responses elicited by candidate human immunodeficiency virus type 1 vaccines. J Virol 79, 10103–10107 (2005).

63. Mishra N, et al. Broadly neutralizing plasma antibodies effective against autologous circulating viruses in infants with multivariant HIV-1 infection. Nat Commun 11, 4409 (2020).

64. Metsalu T, Vilo J. ClustVis: a web tool for visualizing clustering of multivariate data using Principal Component Analysis and heatmap. Nucleic Acids Res 43, W566–570 (2015).

65. Firth HV, et al. DECIPHER: Database of Chromosomal Imbalance and Phenotype in Humans Using Ensembl Resources. Am J Hum Genet 84, 524–533 (2009).

66. R, Core, Team. R: A language and environment for statistical computing. (2017).

67. Wickham H. ggplot2: Elegant Graphics for Data Analysis. Springer-Verlag (2016).

68. Mellors JW, et al. Plasma viral load and CD4+ lymphocytes as prognostic markers of HIV-1 infection. Ann Intern Med 126, 946–954 (1997).

69. Walker LM, et al. Broad neutralization coverage of HIV by multiple highly potent antibodies. Nature 477, 466–470 (2011).

70. Trkola A, et al. Human monoclonal antibody 2G12 defines a distinctive neutralization epitope on the gp120 glycoprotein of human immunodeficiency virus type 1. J Virol 70, 1100–1108 (1996).

71. Brunel FM, et al. Structure-function analysis of the epitope for 4E10, a broadly neutralizing human immunodeficiency virus type 1 antibody. J Virol 80, 1680–1687 (2006).

72. Barroso H, et al. Evolutionary and structural features of the C2, V3 and C3 envelope regions underlying the differences in HIV-1 and HIV-2 biology and infection. PLoS One 6, e14548 (2011).

73. Mabvakure BM, et al. Positive Selection at Key Residues in the HIV Envelope Distinguishes Broad and Strain-Specific Plasma Neutralizing Antibodies. J Virol 93, (2019).

74. Dingens AS, Arenz D, Weight H, Overbaugh J, Bloom JD. An Antigenic Atlas of HIV-1 Escape from Broadly Neutralizing Antibodies Distinguishes Functional and Structural Epitopes. Immunity 50, 520–532 e523 (2019).

75. Landais E, Moore PL. Development of broadly neutralizing antibodies in HIV-1 infected elite neutralizers. Retrovirology 15, 61 (2018).

76. Bricault CA, et al. HIV-1 Neutralizing Antibody Signatures and Application to Epitope-Targeted Vaccine Design. Cell Host Microbe 26, 296 (2019).

77. Local, Burden, of, Disease, HIV, Collaborators. Subnational mapping of HIV incidence and mortality among individuals aged 15-49 years in sub-Saharan Africa, 2000-18: a modelling study. Lancet HIV 8, e363–e375 (2021).

78. Sebastiao CS, Morais J, Brito M. Factors Influencing HIV Drug Resistance among Pregnant Women in Luanda, Angola: Findings from a Cross-Sectional Study. Trop Med Infect Dis 6, (2021).

79. Gray ES, et al. The neutralization breadth of HIV-1 develops incrementally over four years and is associated with CD4+ T cell decline and high viral load during acute infection. J Virol 85, 4828–4840 (2011).

80. Moore PL. The Neutralizing Antibody Response to the HIV-1 Env Protein. Curr HIV Res 16, 21–28 (2018).

81. Bhiman JN, et al. Viral variants that initiate and drive maturation of V1V2-directed HIV-1 broadly neutralizing antibodies. Nat Med 21, 1332–1336 (2015).

82. Liao HX, et al. Co-evolution of a broadly neutralizing HIV-1 antibody and founder virus. Nature 496, 469–476 (2013).

83. Doria-Rose NA, et al. Developmental pathway for potent V1V2-directed HIV-neutralizing antibodies. Nature 509, 55–62 (2014).

84. Ndlovu B, et al. Envelope characteristics in individuals who developed neutralizing antibodies targeting different epitopes in HIV-1 subtype C infection. Virology 546, 1–12 (2020).

85. Beirnaert E, et al. Identification and characterization of sera from HIV-infected individuals with broad cross-neutralizing activity against group M (env clade A-H) and group O primary HIV-1 isolates. J Med Virol 62, 14–24 (2000).

86. Kouyos RD, et al. Tracing HIV-1 strains that imprint broadly neutralizing antibody responses. Nature 561, 406–410 (2018).

87. Sather DN, et al. Factors associated with the development of cross-reactive neutralizing antibodies during human immunodeficiency virus type 1 infection. J Virol 83, 757–769 (2009).

88. Moir S, Fauci AS. B-cell responses to HIV infection. Immunol Rev 275, 33–48 (2017).

89. Badura R, et al. Early ART in Acute HIV-1 Infection: Impact on the B-Cell Compartment. Front Cell Infect Microbiol 10, 347 (2020).

90. Moir S, et al. B cells in early and chronic HIV infection: evidence for preservation of immune function associated with early initiation of antiretroviral therapy. Blood 116, 5571–5579 (2010).

91. Gach JS, et al. HIV-1 specific antibody titers and neutralization among chronically infected patients on long-term suppressive antiretroviral therapy (ART): a cross-sectional study. PLoS One 9, e85371 (2014).

92. Kong R, et al. Broad and potent neutralizing antibody responses elicited in natural HIV-2 infection. J Virol 86, 947–960 (2012).

93. Rodriguez SK, et al. Comparison of heterologous neutralizing antibody responses of human immunodeficiency virus type 1 (HIV-1)- and HIV-2-infected Senegalese patients: distinct patterns of breadth and magnitude distinguish HIV-1 and HIV-2 infections. J Virol 81, 5331–5338 (2007).

94. Rocha C, et al. Potency of HIV-2-specific antibodies increase in direct association with loss of memory B cells. AIDS 31, 2431–2433 (2017).

95. Bai H, Li Y, Michael NL, Robb ML, Rolland M. The breadth of HIV-1 neutralizing antibodies depends on the conservation of key sites in their epitopes. PLoS Comput Biol 15, e1007056 (2019).

96. Rademeyer C, et al. Genetic characteristics of HIV-1 subtype C envelopes inducing cross-neutralizing antibodies. Virology 368, 172–181 (2007).

97. Kumar S, et al. An HIV-1 Broadly Neutralizing Antibody from a Clade C-Infected Pediatric Elite Neutralizer Potently Neutralizes the Contemporaneous and Autologous Evolving Viruses. J Virol 93, (2019).

98. Cheedarla N, et al. Evolution of Neutralization Response in HIV-1 Subtype C-Infected Individuals Exhibiting Broad Cross-Clade Neutralization of HIV-1 Strains. Front Immunol 9, 618 (2018).

99. Lei L, et al. The HIV-1 Envelope Glycoprotein C3/V4 Region Defines a Prevalent Neutralization Epitope following Immunization. Cell Rep 27, 586–598 e586 (2019).

